# CO2 measurements in instrumental and vocal closed room settings as a risk reducing measure for a Coronavirus infection

**DOI:** 10.1101/2020.10.26.20218354

**Authors:** Manfred Nusseck, Bernhard Richter, Ludwig Holtmeier, Dominik Skala, Claudia Spahn

## Abstract

Contaminated aerosols in room air are one of the transmission routes of the coronavirus. The amount of contaminated aerosols in the room seems to play an important role for the infection risk. In rooms without technical air refreshing systems, the aerosol concentration can be reduced with simple natural ventilation activity. Instrumental and vocal lessons and rehearsals take place in closed indoor rooms. Therefore it is important to optimize the necessary ventilation activity in order to keep the infection risk for musicians low. Therefore, knowledge about the maximum duration of the lesson or rehearsal for ventilation intervals are necessary. In this study, carbon dioxide concentration (CO2) as an indicator of the indoor air quality (IAQ) was measured during 47 music lessons and rehearsals at a university of music including 141 persons. From these measurements, the air exchange rates of the rooms and the CO2 emission rates per person were extracted. The results show that the CO2 emission in musical activities can be assigned to light and moderate activities between 28 l/h and 39 l/h. Wind instruments had the highest CO2 emissions. Singers showed low CO2 emission rates comparable to the control group which only spoke and listened. Recommendations for the frequency of ventilation breaks were derived from empirical data and allow for an individual risk assessment of instrumental and vocal lessons and rehearsals depending on room size and number of musicians.

## INTRODUCTION

The COVID-19 pandemic led to severe restrictions in everyday social and occupational activities. One of the most relevant modes of transmission of the coronavirus in human-to-human interaction is through contaminated aerosols.[1] These aerosols are exhaled by an infected person and then inhaled by another person.[2] Reduction of aerosol transmission can be accomplished by performing regular effective ventilation activities in indoor rooms. Another way of transmission of SARS-CoV-2 are larger particles, so called droplets. Plausible protections against infections via droplets and aerosols can be accomplished by considering social distancing and wearing a mouth and nose mask.[3, 4]

### Aerosols

Airborne aerosols are droplets of different sizes interspersing with the surrounded air and with small sizes of less than 5µm diameter can remain in the air and freely drift away by air currents to far distances [1]. Aerosols originate of respiratory secretion through breathing, talking, coughing, sneezing, and singing. If a person is infected with SARS-CoV-2, the exhaled aerosols may contain the virus.

Especially in indoor rooms, the aerosol concentration may increase rapidly and with it the potential risk of infection. Thus, closed rooms without technical air ventilation systems may become a serious risk environment especially during autumn and winter time. Kriegel & Hartmann [5] designed a theoretical model of aerosol growth assuming a similar spreading and infection procedure of SARS-CoV-2 as in comparison to the influenza virus.[6] Their calculations are based on the assumption that the inhalation of 3000 virus – one virus on one aerosol – may lead to an infection with SARS-CoV-2. From this, they derive that, for example, in a room with one infectious person doing light work with a floor size of 25m^2^, a ceiling height of 3m – that means a room volume of 75m^3^ – and a low air exchange rate the critical level of aerosol concentration for an infection with SARS-CoV-2 is reached after about one hour.[5]

To keep the concentration of contaminated aerosols in rooms at a low level, regular ventilation activity has been shown to effectively reduce the infection risk.[2, 7, 8] Hence, it is important to know, when a room in which a number of persons are doing certain activities needs to be ventilated before a critical concentration has been reached.

### CO2 as indicator for indoor air quality

The concept of indoor air quality (IAQ) is widely used and Carbon dioxide (CO2) is a reasonable reference of the IAQ in rooms.[9] Several studies considered the IAQ by measuring the CO2 concentration especially in schools.[10, 11] There are also studies showing correlations of the CO2 concentration with present air pollutants.[12] For SARS-CoV-2, according to the current scientific state of knowledge there are no verified direct correlations between the CO2 concentration and the amount of aerosols especially of virus-containing aerosols in indoor room existent. Hartmann & Kriegel [13] tried to mathematically model the possibility of being infected by SARS-CoV-2 in relation to the CO2 concentration. By using specific approximations for the model, they imply a theoretical connection between CO2 and aerosol emission. However, as a control of the IAQ, CO2 measurements can be used as an indicator of when a ventilation activity should be necessary and with the ventilation, the aerosol loading will similarly be reduced. [8, 14]

The CO2 concentration is measured in ppm (parts-per-million) and defines the number of carbon dioxide particles in a given number of particles in the air. The global atmosphere currently contains about 400ppm of CO2. There are seasonal dependencies regarding the differences in temperature and air humidity.[15] Higher levels of CO2 above 2000ppm increase the frequency of nausea and drowsiness. [16] Standards such as the ASHRAE (American Society of Heating, Refrigerating and Air-Conditioning Engineers, [17]) and the EN13779 (European standard for mechanical ventilation and air conditioning, [18]) suggest a maximum indoor CO2 level of 1000 ppm. This level has been defined as the threshold of good air quality by Pettenkofer in 1858, already. The EN13779 classifies a CO2 concentration below 800ppm as high indoor air quality (IDA 1) and between 800ppm and 1000ppm as medium indoor air quality (IDA 2). The German ASA A3.6 [19] recommends keeping the level of indoor CO2 below 1000ppm.

The increase of the CO2 concentration follows an exponential convergence where the saturation is given by the emitting CO2 source and the air change rate in the room.[20] In rooms with technical air ventilation systems the saturation can be reached quickly at a rather low concentration level. In closed rooms without technical air ventilation systems the saturation point can be very high and the CO2 concentration increases approximately linear.

As the main CO2 source is the human respiration, the CO2 emission rate depends upon the activity of the person(s) in a room.[9] The more physical activity, the more CO2 is been emitted.[20] The amount of exhaled CO2 is measured in l/h (liter per hour). The general value of emitted CO2 of an adult person doing a light activity such as sitting at a desk is about 20 l/h.[9] For a moderate activity, such as standing and doing moderate work, the value is about 34 l/h [7] and for strenuous activities such as sports (e.g. calisthenics with moderate effort) and physically demanding work, the CO2 emission can go above 70 l/h.[9]

According to the BGN [7], the level of 1000ppm is reached after about one hour, if two persons are doing light work in a room of 75m^3^ without air ventilation system. Kriegel & Hartmann [5] calculated for the same room that an infectious person increases the aerosol concentration within the same time to a certain risk level of infection. Both thresholds are rather close together. In order to avoid a possible risk of infection, however, we propose to address a threshold of CO2 concentration below 1000ppm as advised by the ASA A3.6.[19] In accordance with other commission of experts, we suggest to use the first air quality level of 800ppm (IDA-1) as the threshold at which ventilation especially during musical activities should take place.

Evaluating the IAQ by measuring the CO2 concentration has been shown to be a reliable indicator for recommendations of effective ventilation.[21, 22] With a technical air ventilation system, the IAQ can be mechanically controlled. Without such system a natural ventilation by opening windows and doors is necessary. The natural ventilation distinguishes between a rapid and a permanent air exchange. For the rapid air exchange, a window is widely opened and if possible additionally a second window or a door at the other side of the room too in order to generate a draft. If a window is slightly opened (tilted) for a longer time, a permanent natural ventilation is established and generates a constant air exchange. With this ventilation technique, a saturation of the CO2 concentration at a low level can be accomplished. However, in cold seasons, this ventilation leads to drastic energy losses. It is therefore recommended to use rapid ventilation procedures in regular intervals.[7, 8]

### Musical settings

Regarding the optimal protection of musicians against Coronavirus infection, there are important risk assessments available.[23, 24] Those risk assessments provide a number of specific recommendations for musicians to reduce the risk of infection during playing and practicing. Within a range of measures, Spahn & Richter [23, 24] considered the use of measuring the CO2 concentration as plausible indicator of IAQ and thus for the infection risk.

Wind instruments and singing have been considered to have high aerosol emission of SARS-CoV-2 as this transmission is mainly produced by exhalation. There are just a few studies regarding the aerosol generation in playing musical instruments. For singing, the aerosol production was found to be on average three times higher than for speaking.[25, 26] In contrast, Gregson et al. [27] found no difference in aerosol production between singing and speaking at a low vocal intensity of 50-60dB. However, they found a relation between the aerosol production and the vocal intensity with more aerosol generation at higher vocal intensities. Only at very high vocal intensity (90-100dB) did they find a difference between singing and speaking with higher aerosol production during singing.

For wind instruments, He et al. [28] investigated the aerosol production for ten different woodwind and brass instruments. Only one instrument, the tuba, were found to be at a low risk level regarding the aerosol emission. Three instruments were at a high risk level including the trombone, the oboe and the trumpet.

However, at present, there are no reliable values for the CO2 emission regarding the activity of singing and playing different instruments.

### Aim of the study

Musical settings typically include instrumental and vocal lessons and rehearsals that take place in closed rooms with at least two persons. Generally, there are settings of one-to-one lessons and rehearsals with two or more musicians. A personal interaction between people, teachers and students, is indispensable. Therefore, to keep the infection risk low in such settings, it is important to know how long a lesson or rehearsal should last before being interrupted for a necessary ventilation break. From measuring the CO2 concentration recommendations for the duration of the lesson or rehearsal and for necessary ventilation cycles can be developed. Furthermore, it was aimed to classify the Co2 emission rate of musical activities compared to light, moderate and strenuous activities.

## METHODS

### Participants and settings

The CO2 emission was recorded in 47 instrumental and vocal scenarios with a total of 141 persons. The measurements took place in the University of Music Freiburg in the commonly used rooms for instrumental and vocal lessons and rehearsals. No personal data were collected from the players, except for the instrument played. All participants have a high level of musical expertise. The CO2 sensor was placed in the room after verbal agreement of the players was given.

In a one-to-one lesson, the student usually plays the instrument or sings and the teacher is listening. Between the playing, both student and teachers are talking and discussing. Sometimes the teacher plays or sings along with the student. In rehearsal situations, players are practicing along with other players. Generally, the musicians play together simultaneously and in playing breaks, they also talk and discuss.

For the analysis, specific activity groups were formed (Table 1). According to the findings of higher aerosol productions in singing [25, 26] and for wind instruments [28], the singers and the brass players were distributed into the “vocal” and the “brass” group respectively. The woodwind instruments were divided into the “low pressure” group, containing flute and recorder, and the “high pressure” group with clarinet, saxophone and oboe. As a non-wind instrument, the strings were put into an individual “strings” group. Further non-wind instruments were assigned to the “others” group. This group contains the piano and the harp as well as percussionists playing moderately the timpani and other percussion instruments. In some settings, a player was accompanied by a different instrument, e.g. a flute and a piano player. Such settings constitute the “mixed instruments” group. As a control group, seminar situations in which people spoke and listened were additionally measured.

**Table 1:** Distribution of the different activity groups with the amount of settings and players. The percentage regards the number of persons.

|  | No. of settings | No. of Persons | Percent of persons |
| --- | --- | --- | --- |
| **Vocal** | 5 | 11 | 7.8% |
| **Brass** | 4 | 12 | 8.5% |
| **Woodwind (low pressure)** | 9 | 25 | 17.7% |
| **Woodwind (high pressure)** | 5 | 14 | 9.9% |
| **Strings** | 9 | 30 | 21.3% |
| **Mixed instruments** | 6 | 14 | 9.9% |
| **Others (i.e. piano, harp, percussion)** | 4 | 8 | 5.7% |
| **Control (speaking and listening)** | 5 | 27 | 19.2% |
| **Total** | 47 | 141 | 100% |

The rooms in which the settings took place had different sizes and their volume ranged between 56m^3^ and 216m^3^ and the height of the rooms varied between 3m and 5m. The mean room volume was 118.8m^3^ (SD = 45.1m^3^). No significant difference was found in the mean room size across the instrumental groups. The mean room volume per person was 44.2m^3^ (SD = 17.2m^3^) with a range between 13.8m^3^ and 108.0m^3^.

### Measuring device and study design

For the measurement of the CO2 concentration the AIRCONTROL 5000 from TFA Dostmann (Germany) was used. The CO2 sensor assesses temperature (in °C), air humidity (in %) and CO2 value in ppm. The CO2 measuring limit goes up to 5000ppm. The values were averaged across 5 second intervals. With a data logger, all measures were saved with a time stamp in a CSV file. The data were continually stored on an external SD card.

As recommended from the literature (see [29]), the CO2 devices were installed on top of a portable stand 110-130cm above the floor. In some rooms the device was mounted on a tripod of about 30cm and was put on a table in the rooms (about 70cm height). To reduce disturbance and diffusion interactions of air movements within the rooms, a distance of 60cm to a wall and a window was maintained. Furthermore, the devices had a distance of at least 2.0m from any occupant to avoid local exhalation effects on the CO2 values.

At the beginning of the instrumental and vocal lesson or rehearsal the CO2 sensor was placed in the room accordingly. The room was completely ventilated before a lesson or rehearsal with open windows and doors to get a low CO2 environment at 400-500 ppm. Windows and doors were closed during lessons or rehearsals and only the players stayed in the room. The measurements during lessons and rehearsals were at least 20 minutes. When the CO2 value during lessons or rehearsals reached 800 ppm, all players left the room and the windows and the doors were opened for a rapid air exchange (boost ventilation). The time was measured, until a value of 400 ppm was reached again.

### Data analysis

For calculating the air exchange rates of the rooms and the CO2 emission rates per person, a standard mathematical model was used.[22, 30] Eq. (1) provides the CO2 concentration after a time t taking into account a total balance of the indoor CO2 distribution

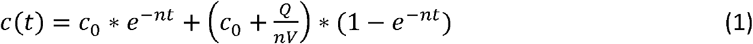

where, c(t) is the monitored CO2 concentration (in ppm) at time t, c_0_ is the CO2 concentration at time t=0, V is the room volume in m^3^, Q is the CO2 emission rate from the source in l/h and n is the air exchange rate in 1/h.

By using Matlab (R2020a), an exponential curve fitting of Eq. (1) was used for the measured CO2 data in every scenario. Such modelling of collected CO2 concentrations has been shown to be an effective method for evaluating indoor air parameters.[21] The equation solver calculated best fitting estimations of the air exchange rate n, the CO2 emission rate Q and the CO2 concentration at time t=0 (c_0_). From the estimated total CO2 emission rate Q the mean CO2 emission rate of the occupants in each setting was calculated by dividing Q by the number of persons.

All parameter values are reported descriptively with mean and standard deviation of the mean (SD). For the analysis of main effects between the activity groups, an analysis of variance (ANOVA) was used. The post-hoc comparisons of the mean values between the activity groups were performed with the Bonferroni correction. Correlations were calculated by bivariate correlations and Pearson’s r is been reported. The statistical analyses were conducted with SPSS (Version 27, Armonk, NY: IBM Corp.). The level of significance was set to p = 0.05.

## RESULTS

The mean measuring results by activity group are listed in Table 2. The mean values across all settings were for the temperature 24.8°C (SD = 1.9°C), the air humidity 43.9% (SD = 4.2%) and for the air exchange rates 0.49 1/h (SD = 0.22 1/h). The mean CO2 emission rate per person was 33.1 l/h (SD = 5.8 l/h). There were no significant correlations between these parameters.

**Table 2:**
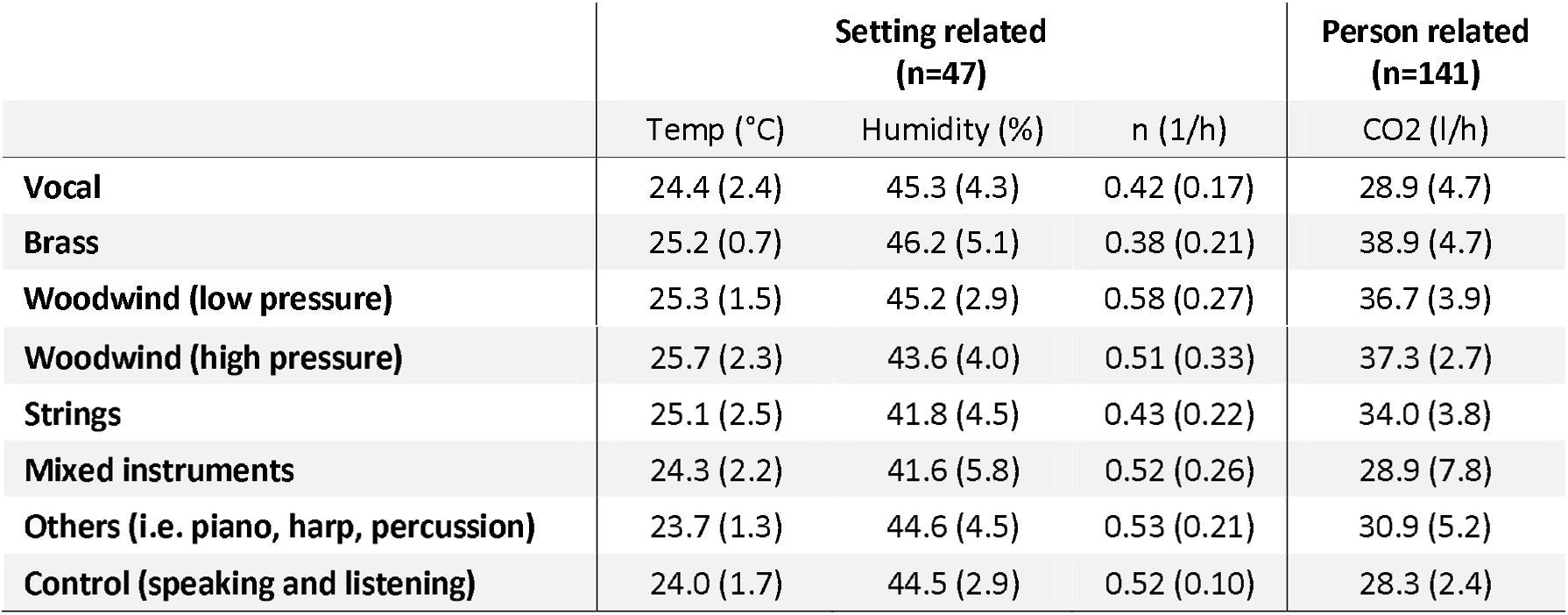
Mean values of the room temperature, the air humidity, the estimated air exchange rate (n) in the setting rooms and the estimated CO2 emission per person by activity group (In brackets: standard deviation of the mean)

The temperature, the humidity and the air exchange rates showed no significant differences across the activity groups. A significant main effect was found for the mean CO2 emission per person between the activity groups (F(7,133)=15.965, p < .001). Figure 1 shows the assignment of different musical activities to light and moderate activities.

**Figure 1:**
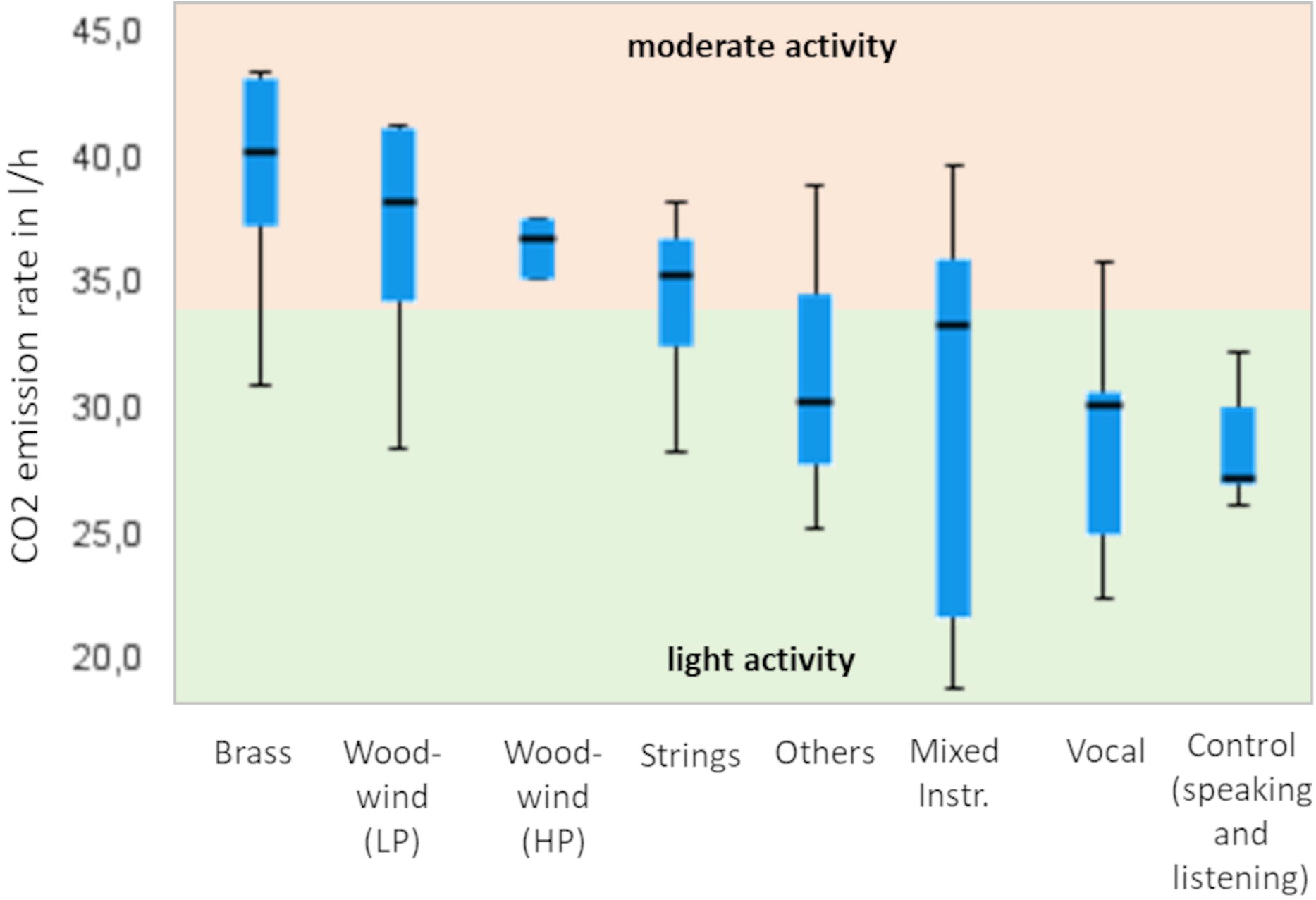
Estimated CO2 emission per person by activity group (LP: low pressure, HP: high pressure, Others: piano, harp, percussion) sorted by the emission rates (green: light activity; orange: moderate activity [7])

There were significant differences in the CO2 emission rates of different musical activities. With a post-hoc pairwise comparison, significant differences in the mean CO2 emission rates were found between the vocal group and the brass group (p < .001), the woodwind low pressure group (p < .001), the woodwind high pressure group (p < .001) and the strings group (p = .019) with lower values in the vocal group. The brass group was also significantly higher in the mean CO2 emission rate than the strings group (p = .035), the mixed instruments group (p < .001), the others group (p = .002) and the control group (p < .001). The woodwind low pressure group showed higher mean rates compared to the mixed instruments group (p < .001) and the control group (p < .001). Similarly, the CO2 emission rates of the woodwind high pressure group were significantly higher than in the mixed instruments group (p < .001), the others group (p = .033) and the control group (p < .001). The strings group were significantly higher in CO2 emission rates than the control group (p < .001).

A significant medium correlation was found between the room volume and the CO2 emission rate per person (r = .44; p = 0.002). The larger the room was, the higher was the mean CO2 emission. The correlation between the air exchange rate and the room size was not significant.

During the rapid air ventilation break, the CO2 concentration went down to a regular level of about 400-500ppm within 10 minutes. However, smaller rooms showed to have a slightly quicker ventilation effect than larger rooms.

The estimated coefficients of the CO2 emission rate per person for each activity group (Table 2) were used to draw theoretical serial data curves with Eq. (1). According to the findings, the air exchange rate n was set to 0.5 1/h. For the room, a room size of 75m^3^ in volume (25m^2^ floor size and 3m ceiling height) was used. Additionally, the room was fictionally filled with two musicians. Figure 2 provides the individual curves of the CO2 dispersal in the room over time.

**Figure 2:**
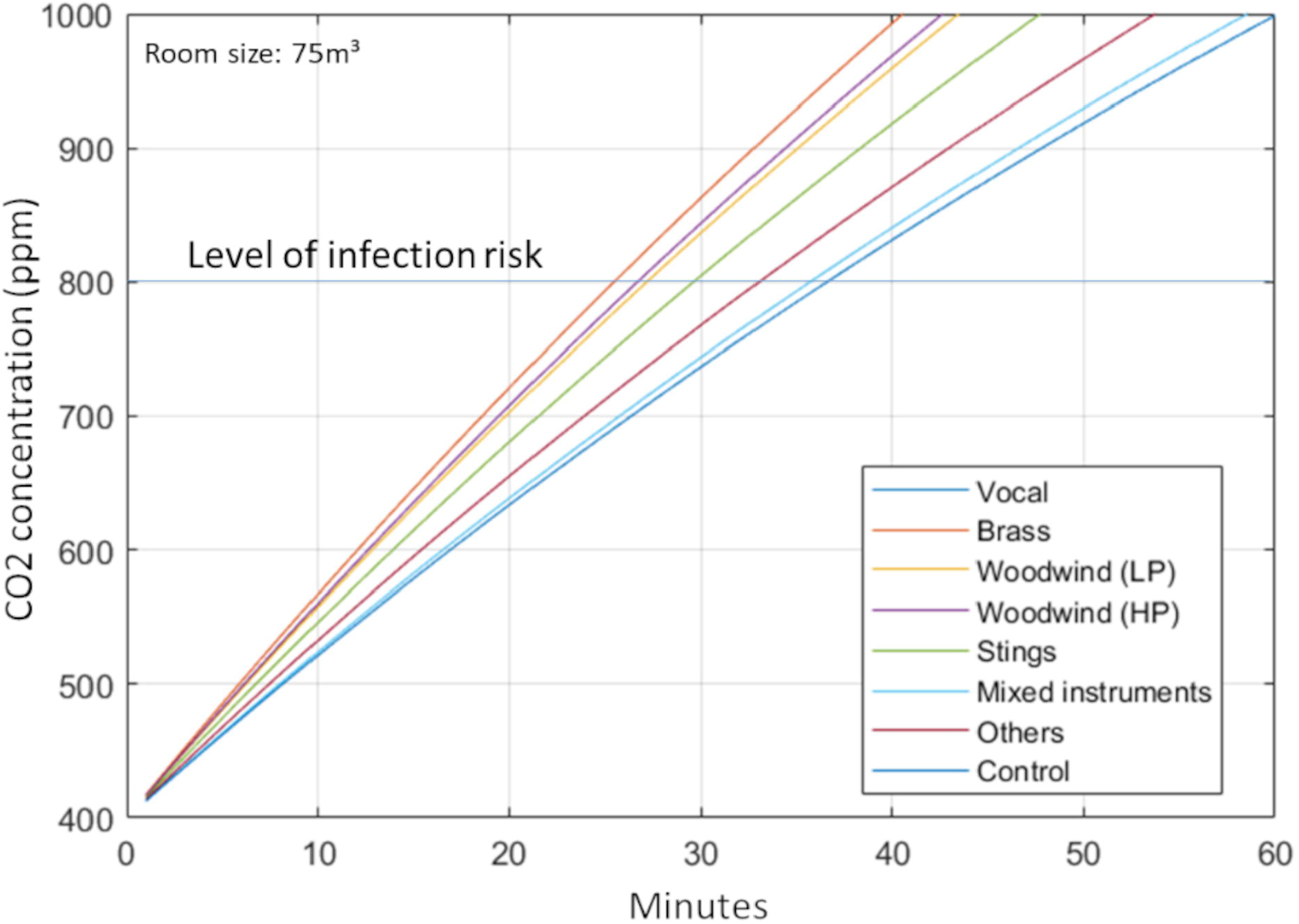
Theoretical curves regarding Eq. (1) for the different activity groups for a room with 75m^3^, an air exchange rate of 0.5 1/h and with two occupants.

The duration of the lesson till the CO2 concentration reaches the critical level of 800ppm ranges between 25 minutes for the brass group and 36 minutes for the vocal and the control group. In a room of 100m^3^ the range of duration increases up to 35 minutes for the brass group and 53 minutes for the vocal and the control group.

## DISCUSSION

In this study, different real settings of instrumental and vocal lessons and rehearsals were investigated for the indoor CO2 emission. Such lessons and rehearsals take place in closed rooms with two or more persons and generally without any kind of technical air ventilation system. Therefore, when the CO2 concentration in the room reaches a critical level of 800ppm a natural ventilation activity is been recommended. This can keep the concentration of possibly contaminated aerosols with SARS-CoV-2 low and effectively reduce the risk of infection. With the empirical data of the CO2 measurements, recommendations for length of stay and ventilation breaks in instrumental and vocal lessons and rehearsals with two musicians are developed.

### CO2 emission rates

An important goal of this study was to collect the CO2 emission rates while singing and playing instruments and to compare them to other activities. The mean estimated CO2 emission rate per person across all scenarios with 33 l/h was slightly lower than the level of moderate activities of 34 l/h.[7] However, the difference between the activity groups was about 10 l/h. The brass and the woodwind groups were above 34 l/h and the vocal, the others, the mixed instruments and the control group were below 34 l/h. The strings group showed to have exactly the value of moderate activities. This finding indicates that different musical activities need to be addressed specifically considering the air quality with regards to the room.

The results of the CO2 emission rates showed that the brass and the woodwind instruments had higher CO2 exposure compared to the other instruments and the control group. In a similar way, He et al. (2020) found that the aerosol production of brass and woodwind instruments were also at high levels. For these instruments, it can be assumed that the CO2 emission and the aerosol production may have a direct connection and are at a high level of exposure. Interestingly, the recorder and the flute who are associated with higher levels of air release during sound production than in the other wind instruments showed no difference in the CO2 emission rates to the high pressure woodwind instruments. However, wind instruments need to be considered more seriously regarding ventilation breaks than other instruments.

The mixed instruments group showed a large variation in the CO2 emission rate. As this group contained different instruments, this indicates that a CO2 emission rate of one person in the room could not be derived by using the mean emission rate. If a brass player and a piano player were in the room, it would be more sufficient to use the individual CO2 emission rate for the brass and the others, i.e. piano. More research is needed to investigate such mixed settings and to get more details on the CO2 emission rates of the individual instruments in the mixed group.

The mean CO2 emission rates between the control group of speaking and listening and the vocal group were not significantly different. Regarding the CO2 emissions, both activities seem to have similar production rates. If we assume a relation between aerosol and CO2 emission this goes in line with the findings of Gregson et al. [27] when considering a medium vocal intensity during the lesson and rehearsal. But even if at first glance our results seem to be in contrast with the results of Mürbe et al.[25] we have to keep in mind that there is no evident correlation between aerosol and CO2 concentration in room air. If the aerosol emission is higher for singing than for speaking, for instance at higher vocal intensity levels, the lesson or rehearsal duration for a vocal activity should be considered to be shorter than the measured results recommend.

The differences between the activity groups in the CO2 emission rates indicates, that a general recommendation for instance of 45 minutes for all lessons is not plausible. The duration of lessons and rehearsals should regard the specific instrumental emission rates. The simulated CO2 exposure in a fictional room with 75m^3^ showed a difference of 11 minutes and with 100m^3^ even about 20 minutes in duration between the extreme groups (Figure 2). To provide a certain length for the lesson or rehearsal besides the specific CO2 emission rate the room size and the number of persons should be taken into account. In addition, the near region of infection risk with heavy droplets must also be considered by taking specific distancing for singing and playing instruments into account.[23, 24, 31]

The correlation between the room volume and the CO2 emission rates per person is quite puzzling. It could be explained by the assumption that in larger rooms the musicians play unconsciously with more intensity than in a small room. However, more research is needed to investigate this relation.

An important parameter for the indoor CO2 dispersal is the air exchange rate. The average estimated air exchange rate in the rooms that were measured in this study, was about 0.5 1/h. This value is in the standard range for closed indoor rooms.[32] Compared to rooms with technical air ventilation systems and an air exchange rate of 5-6 1/h closed indoor rooms need regular ventilation breaks as with people in these rooms the CO2 and thus the aerosol concentration increases quickly.

### Ventilation

For the ventilation, the natural rapid air ventilation procedure by opening a window and a door at opposite sides of the room was very successful for refreshing the IAQ. This ventilation procedure should therefore be considered as essential activity instead of just tilting one window. It is very effective and saves heating energy especially in cold seasons.[7, 8, 32] In this study, a ventilation break of 10 minutes was sufficient to completely set the CO2 concentration in the room to a level of the outside concentration. However, the duration of the ventilation depends on the temperature difference between indoor and outside. At higher temperature differences the length of the ventilation procedure can be shorter. At least, a minimum of 5 minutes should be considered and 10 minutes are highly recommended. Since this study has been performed in summer, more research on the ventilation duration especially in the winter season is needed.

### Summary

In summary, this is a first study of evaluating the CO2 exposure in real musical situations. The findings confirm the use of measuring the CO2 concentration to provide recommendations of ventilation breaks. However, the drawn recommendations are only valid if also other precautions as distancing, wearing masks etc. will be considered. Therefore, during instrumental and vocal lessons and rehearsals all risk assessments should be equally respected.

## Data Availability

The data can be requested by the coresponding author.

